# Early prone positioning does not improve the outcome of patients with mild pneumonia due to SARS-CoV-2: results from an open-label, randomized controlled trial (the EPCoT Study)

**DOI:** 10.1101/2022.11.12.22282252

**Authors:** Miriam Fezzi, Laura Antolini, Alessandro Soria, Luca Bisi, Francesca Iannuzzi, Francesca Sabbatini, Marianna Rossi, Silvia Limonta, Alban Rugova, Paola Columpsi, Nicola Squillace, Sergio Foresti, Ester Pollastri, Maria Grazia Valsecchi, Guglielmo Marco Migliorino, Paolo Bonfanti, Giuseppe Lapadula, EPCoT Study Group

**Affiliations:** School of Medicine and Surgery, University of Milano-Bicocca, Milan, Italy; Bicocca Bioinformatics Biostatistics and Bioimaging Center - B4, University of Milano–Bicocca, Milan, Italy; Clinic of Infectious Diseases, San Gerardo Hospital, ASST Monza, Monza, Italy

## Abstract

**Background:** Prone positioning (PP) is routinely used among patients with COVID-19 requiring mechanical ventilation. However, its utility among spontaneously breathing patients is still debated.

**Methods:** In an open-label randomized controlled trial, we enrolled patients hospitalized with mild COVID-19 pneumonia, whose PaO2/FiO2 ratio was >200 mmHg and who did not require mechanical ventilation (MV) or non-invasive ventilation (NIV) at hospital admission. Patients were randomized 1:1 to PP on top of standard of care (intervention group) *versus* standard of care only (controls). The primary composite outcome included death, MV, NIV and PaO2/FiO2 <200 mmHg; secondary outcomes were oxygen weaning and hospital discharge.

**Results:** Sixty-one subjects were enrolled, 29 adjudicated to PP and 32 to the control group. By day 28, 11 patients required NIV, 4 MV and 3 died. Overall, 24/61 (39.3%) met the primary outcome. Using an intention-to-treat approach, 15/29 patients in PP group *versus* 9/32 controls met the primary outcome, corresponding to a significantly higher risk of progression among those randomized to PP (HR 2.38 95%CI 1.04-5.43; P=0.040). Using an as-treated approach, which included in the intervention group only patients who maintained PP for ≥3 hours/day, no significant differences were found between the two groups (HR 1.77; 95%CI 0.79-3.94; P=0.165). Also, we did not find any statistically difference in terms of time to oxygen weaning or hospital discharge between study arms, in any of the analyses conducted.

**Conclusions:** We observed no clinical benefit from awake PP among spontaneously breathing patients with COVID-19 pneumonia requiring conventional oxygen therapy.

## BACKGROUND

Prone positioning (PP) is widely recognized as an effective treatment for Acute Respiratory Distress Syndrome (ARDS) requiring mechanical ventilation, due to its beneficial effects on respiratory mechanics, gas exchange and, ultimately, patients’ survival [1-4]. Thus, in the last three years, PP has been extensively used in intensive care units (ICU) to treat Coronavirus Disease 2019 (COVID-19)-related acute hypoxemic respiratory failure requiring mechanical ventilation [5,6]. Concurrently, researchers have started to explore the effects of PP in awake, non-intubated patients with COVID-19. Early observational studies suggested that PP could improve oxygenation in non-intubated patients with COVID-19 [7-9]. Following these preliminary results, awake PP was endorsed by several health care providers worldwide and proposed for hospitalised patients or even suggested for those treated at home or recently discharged [10-13].

However, evidences on durable and clinically meaningful benefits from PP in patients spontaneously breathing are limited and often contradictory [14-20]. We therefore aimed at assessing whether early PP in patients with initial COVID-19 related pneumonia, not requiring invasive or non-invasive (NIV) mechanical ventilation, could improve patients’ outcome.

## METHODS

### Study design

The Early Pronation as Covid Treatment (EPCoT) study is a pragmatic, open-label, monocentric randomised controlled trial conducted on patients admitted to the Infectious Diseases Ward of the “San Gerardo Hospital” (Monza, Italy) between August 15th, 2021, and May 31st, 2022. The study protocol was approved by the Local Independent Ethics Committee before the beginning of the trial and registered on ClinicalTrials.gov (registration number NCT05008380). For each patient, informed consent was obtained prior to enrolment.

### Patients

Patients hospitalised due to COVID-19 were eligible for enrolment if ≥18 years old, had a positive polymerase chain reaction test for SARS-CoV-2 RNA on a respiratory sample within 7 days of the enrolment and presented at least one of the following conditions: (i) radiological evidence of pneumonia or (ii) clinical evidence of respiratory disease, defined as either room air arterial oxygen partial pressure (PaO_2_) <80 mmHg or peripheral saturation (SpO_2_) <94% or need for oxygen supplementation in order to maintain SpO_2_ >93%. Patients were ineligible if they had already undergone PP, if their arterial oxygen partial pressure to fractional inspired oxygen ratio (PaO_2_/FiO_2_) was <200 mmHg, if they required supplementary oxygen by high flow nasal cannula (HFNC) or non-invasive ventilation (NIV) and if they had any contraindications or conditions that could hinder PP (including, but not limited to, unstable fractures, deep venous thrombosis, late pregnancy, altered mental status).

### Randomization and study procedures

Using permuted blocks of variable sizes (4, 6 or 8), participants were randomised 1:1 to awake PP on top of standard of care (intervention group) *versus* standard of care only (control group). The randomization was stratified by symptoms duration before enrolment (≤ or >10 days) and need of oxygen therapy (yes or no). Due to the nature of the intervention, blinding for patients and for the clinical staff was precluded. Study participant allocation was concealed using sealed opaque envelopes, prepared by separate persons not involved in patients’ enrolment and care. Patients allocated to awake PP were encouraged to adopt prone positioning for at least 3 consecutive hours (up to 6 hours according to tolerability) twice a day; patients in the control group were free to adopt and maintain any position during the day.

Patients from both groups received standard of care treatment for COVID-19 pneumonia according to the evidence available at the moment.

### Data collection

Demographic, clinical and anthropometric data were collected at enrolment. Vital signs, type of oxygen support, fraction of inspired oxygen (FiO_2_), clinical status according to the Ordinal Scale for Clinical Improvement (OSCI) from the World Health Organisation [21] and blood gas analysis were recorded at baseline and 1, 3, 7, 14, 21 and 28 days after randomization. Patients discharged before day 28 received additional telephonic follow-up on the 28th day. During follow-up visits, we also assessed the time spent in the prone position in the previous 24 hours, according to patients’ self-reports. Occurrence of adverse events and therapy administered to patients were also recorded. In the event of death, the day and cause of death were recorded.

### Outcome measures

The primary outcome was a combination of death, mechanical ventilation (MV), NIV, or PaO_2_/ FiO_2_ ratio <200 mmHg (whichever came first). Secondary outcomes were time to oxygen weaning (defined as clinical status of 1, 2 or 3 on the OSCI scale), time to hospital discharge, change in clinical status and in PaO_2_/ FiO_2_ ratio during follow-up, and rate of adverse events.

### Statistical analysis

Data were checked for completeness and consistency. Characteristics of the two groups, as defined by the randomization arm, were described using relative frequencies, means and standard deviations, as appropriate. Survival times were calculated starting from the enrolment date for the all the endpoints of interest, but time to hospital discharge, which was calculated starting from the date of hospital admission. The related event indicators were set equal to 1 if observed during the observation period and 0 otherwise. The analyses of the end-points of interest were performed drawing the cumulative incidence / survival curves over time by the Kaplan Meier method. The survival times in the two treatment groups, as defined by the treatment allocated at randomization (intention-to-treat analysis) or by the actual treatment received (as-treated analysis), were compared using the exponential regression model. Multivariable models, adjusted for patients’ age and baseline PaO2/FiO2 ratio, were also conducted.

Patients’ clinical status across time was described using barplot by randomization arm. Changes in PaO2/FiO2 ratio over time were analyzed by a general linear mixed model including interactions between treatment arm and time points.

Assuming a 60% risk of clinical progression to the composite outcome and a 10% loss at follow-up and with the goal of detecting a relative difference of 50% between the intervention and control group (two-sided α of 0.05 and β of 0.8), a sample size of 96 patients (48 for each group) was originally planned. However, due to the reduction in COVID-19 pneumonia incidence and to an overall low enrolment rate, patients’ enrolment was terminated earlier, not reaching the planned sample size. Final follow-up of the last patient enrolled was completed on June 20th, 2022.

All analyses were conducted using Stata (StataCorp, Release 17.0) and used two-sided P values and an α of 0.05.

## RESULTS

### Patients’ characteristics

Sixty-one patients were enrolled and randomized. During the randomization process, two participants were incorrectly randomized as they belonged to the stratum of patients with symptoms for ≤10 days although they had symptoms for 11 and 12 days at the time of randomization. In the end, among the 61 patients enrolled, 29 were adjudicated to PP and 32 to the control group (**Figure 1**).

**Figure 1.**
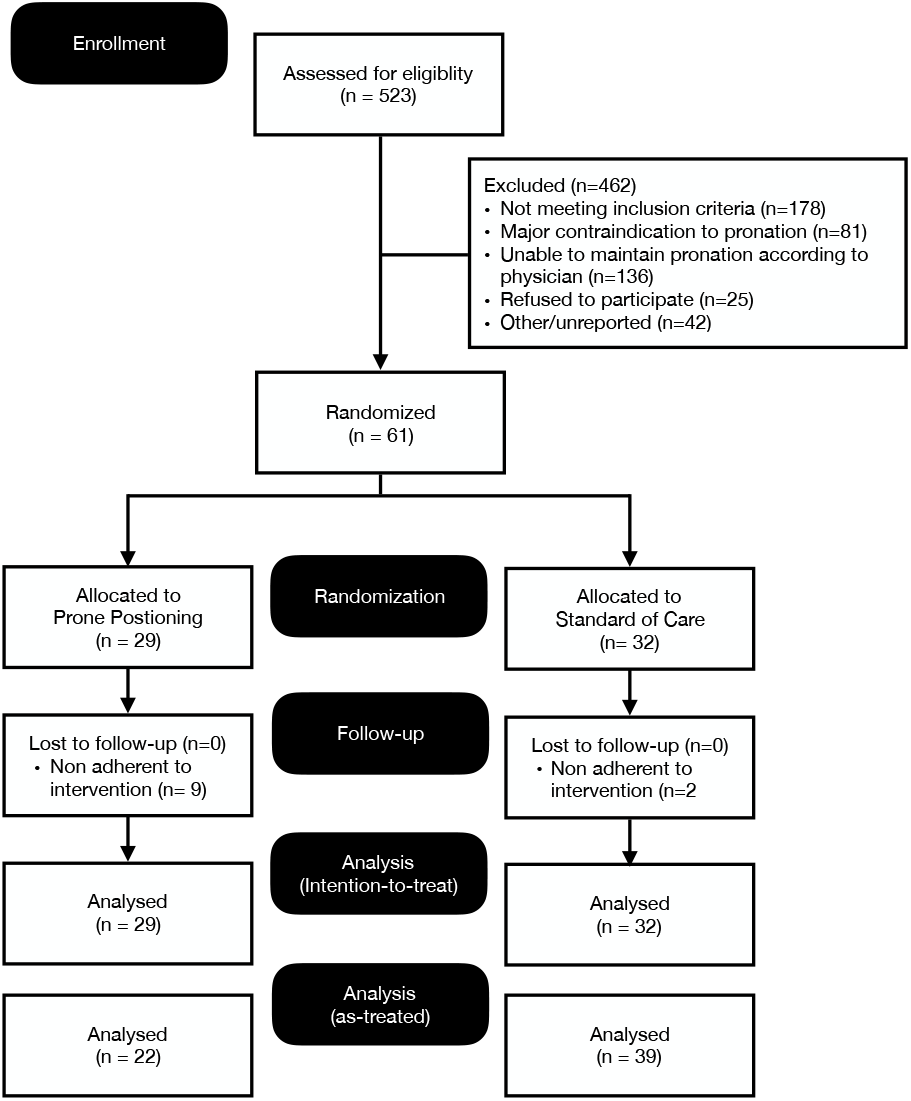
Consort Diagram showing subject disposition in the trial.

The patients enrolled were predominantly male (62.3%) with a mean age of 59.2 (SD 15.7) years. The most common comorbidities were hypertension (29.5%), drug-induced immunosuppression (25.6%) and history of solid or haematological malignancy (16.4% and 11.5%). Half of the patients (50.8%) were vaccinated against COVID-19 with at least one dose. Patients assigned to PP tended to have a slightly more severe disease, as demonstrated by lower PaO_2_/FiO_2_ ratio (280 *versus* 309.8). At the time of enrolment, all patients were on oxygen treatment but one, who required it anyhow later on, during the follow-up. Roughly 89% and 64% of the patients received treatment with corticosteroids (mainly dexamethasone) and remdesivir, respectively. These and other characteristics are shown in **Table 1**.

**Table 1.**
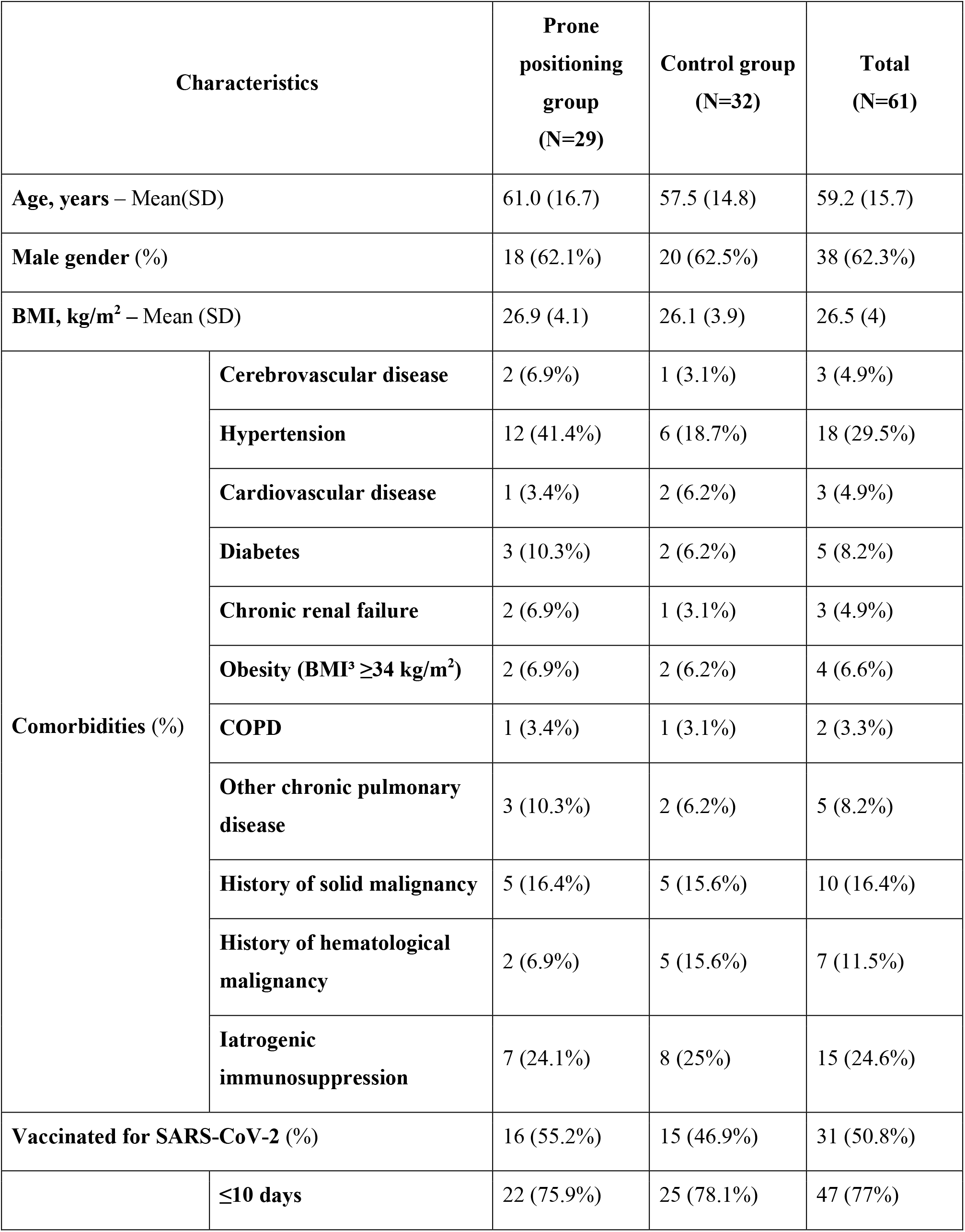
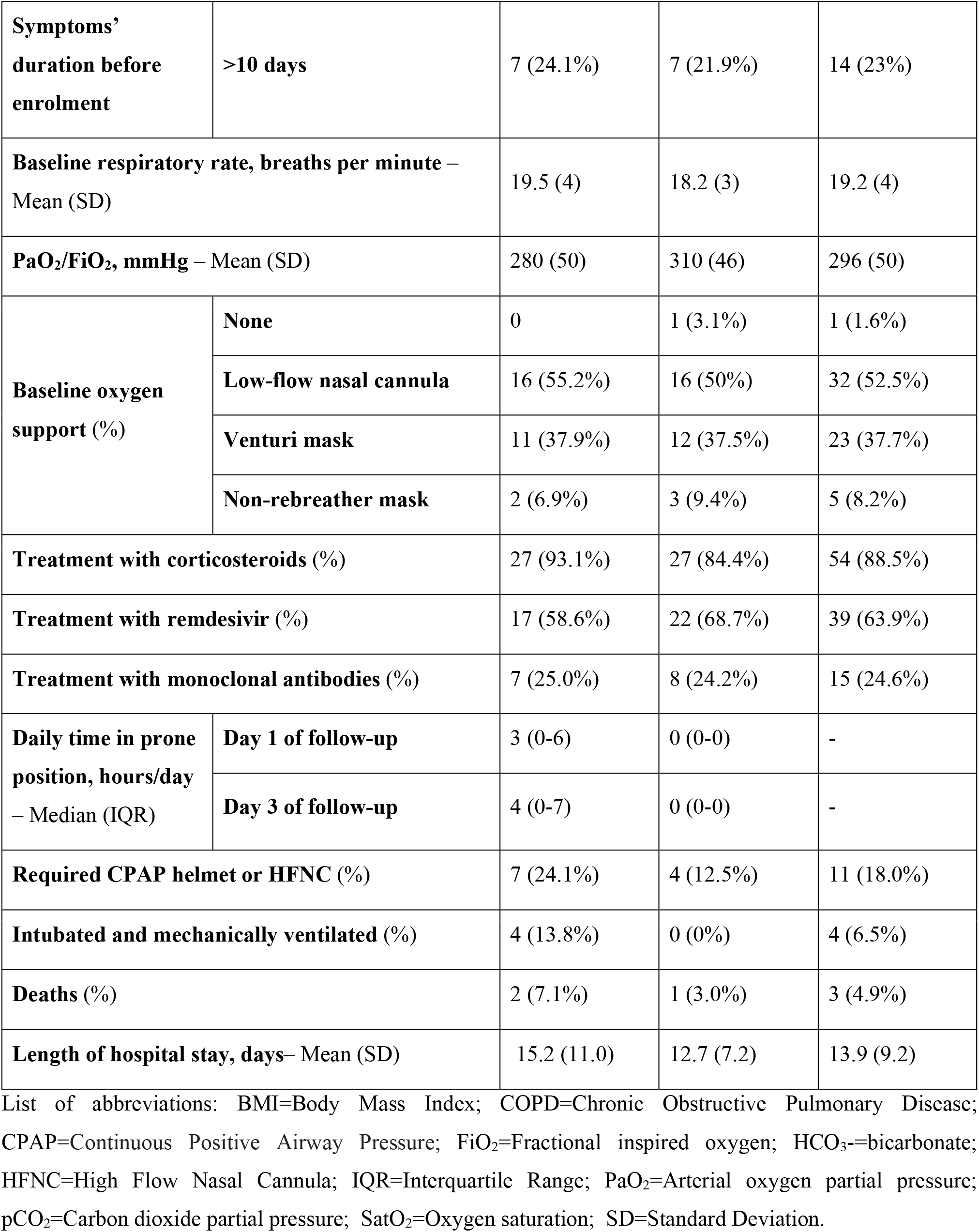
Patient characteristics.

In the intervention group, the median daily time spent in prone positioning on day 1 was 3 hours (IQR 0-6 hours/day), compared to 0 hours/day among the controls; on day 3 we registered a median of 4 hours/day (IQR 0-7) and 0 hours/day, respectively. Nonetheless, suboptimal adherence to the adjudicated treatment was observed in patients randomised to PP, as 44.8% and 40.7% of them maintained pronation for less than 3 hours/day on day 1 and 3, respectively.

### Patients’ outcome and disease progression

By the end of the study, 56 patients (91.8%) were discharged alive, 3 died and 2 were still hospitalised (1 in the Infectious Diseases Ward and 1 in the ICU). During hospitalisation, 11 (18%) patients required non-invasive ventilation with CPAP helmet and 4 (6.5%) orotracheal intubation and mechanical ventilation. The mean length of hospital stay was 15.2 (SD 11.0) days for the prone positioning group and 12.7 (SD 7.2) for the control group.

Figure 2. shows clinical status distribution across the follow-up in the two arms of the study. By day 28, 24/61 (39.3%) patients had reached the primary outcome: 16 because they had a PaO_2_/ FiO_2_ ratio <200 mmHg, 5 because they required NIV and 3 because they were intubated. Using an intention-to-treat approach (**Figure 3A**), the proportion of patients experiencing the primary outcome was 15/29 (51.7%) in the PP group and 9/32 (28.1%) among controls. This resulted in a significantly higher event rate among the intervention group (25.0 [95% CI 15.1-41.5] per 100 person-weeks of observation) than among controls (10.5 [5.5-20.2]), corresponding to a hazard ratio (HR) of 2.38 (95%CI 1.04-5.43; P=0.040). The association between PP and a worse outcome held in a multivariable model adjusted for age and PaO_2_/FiO_2_ ratio at enrolment. In this model, patients randomized to PP still had higher risk of meeting the primary end-point than controls (Adjusted HR 2.35; 95%CI 1.02-5.38; P=0.044), while higher PaO_2_/FiO_2_ ratio was protective (Adj HR 0.4 [0.12-1.34] and 0.27 [0.09-0.79] for PaO_2_/FiO_2_ 250-300 mmHg and >300 mmHg *versus* <250 mmHg, respectively)

Similarly, in an additional analysis only considering clinical end-points (death, endotracheal intubation or NIV), we registered a significant higher event rate in those randomized to PP than in the controls, with a hazard ratio of 4.04 (95%CI 1.28-12.68; P=0.017). Such a significant association held also in the multivariable model, adjusted for the same covariates mentioned above (Adj HR 3.6; 95%CI 1.09-11.83; P=0.034).

**Figure 2.**
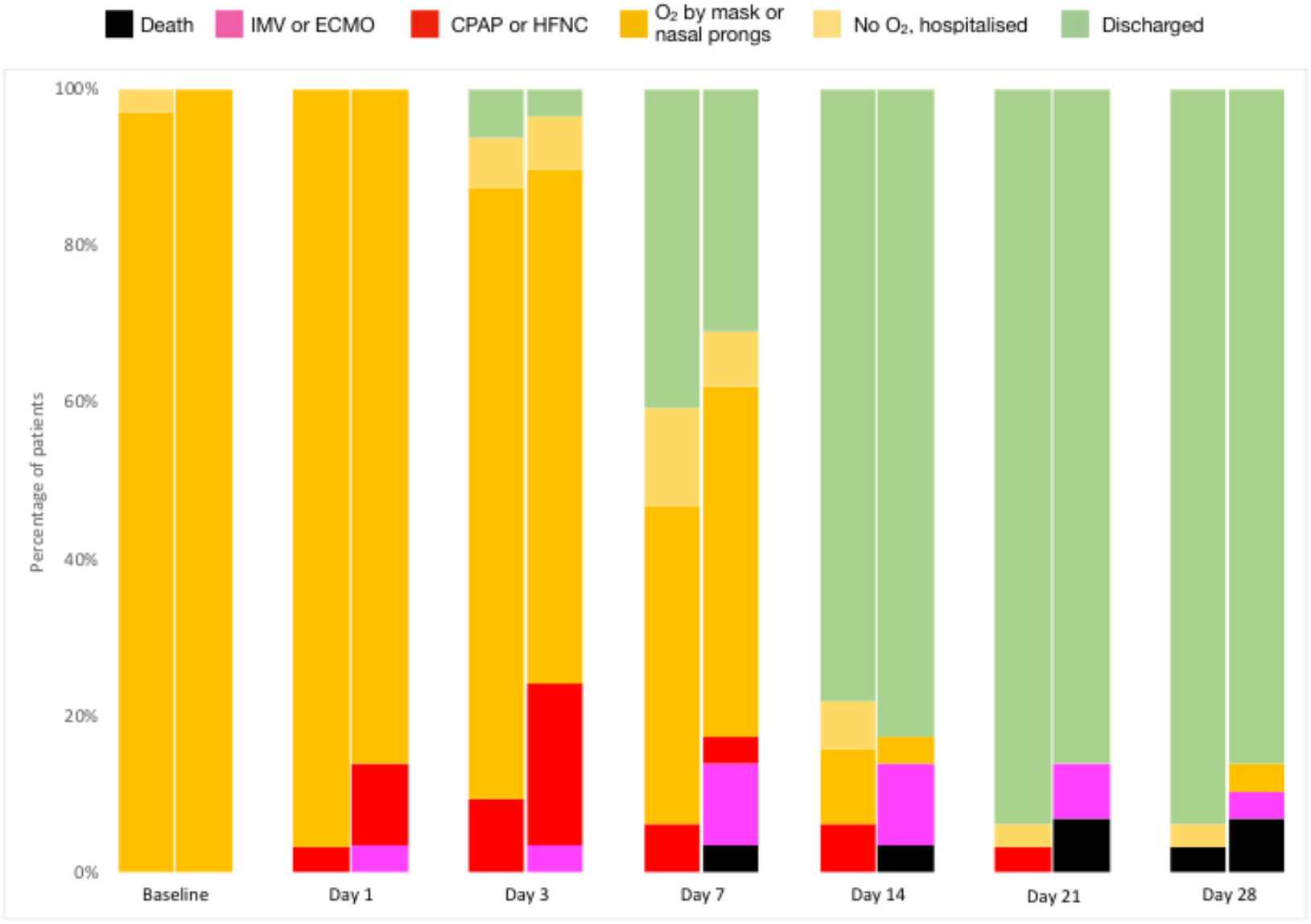
Clinical Status distribution over time, grouped by randomization arm. At each time-point, the left column represents the control arm and the right column represents the awake prone positioning (intervention) arm List of abbreviations: CPAP, continuous positive airway pressure; ECMO, extra-corporeal mechanical oxygenation; HFNC, high-flow nasal cannula; IMV, invasive mechanical ventilation; O_2_, oxygen

**Figure 3.**
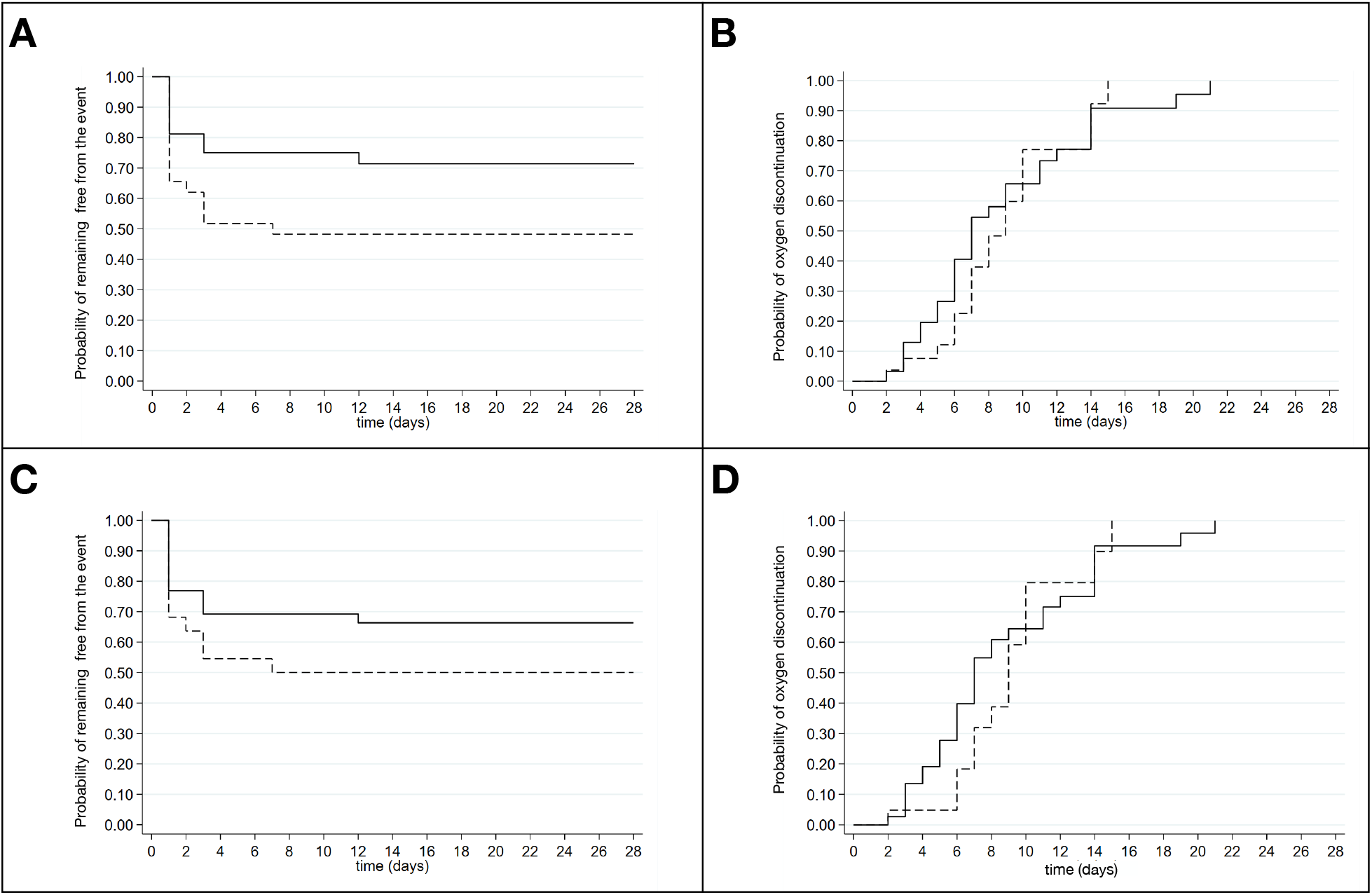
Risk of clinical progression and probability of oxygen weaning by randomization arm (intention-to-treat and as-treated analyses) Panel A: Risk of clinical progression, defined as death, mechanical ventilation, need for high-flow nasal cannula (HFNC) or non-invasive ventilation with continuous positive airway pressure (CPAP). Intention-to-treat analysis; Panel B: Probability of oxygen discontinuation by randomization arm. Intention to treat analysis; Panel C: Risk of clinical progression, defined as death, mechanical ventilation, need for high-flow nasal cannula (HFNC) or non-invasive ventilation with continuous positive airway pressure (CPAP). As-treated analysis; Panel D: Probability of oxygen discontinuation by randomization arm. As-treated analysis. The continuous line represents the control group, the dashed line represents the prone positioning group.

Prone-positioning was not associated either with an accelerated oxygen weaning or hospital discharge. As a matter of fact, as shown in **Figure 3B**, rates of oxygen discontinuation were similar between the two study groups (63.0 [95% CI 39.7-100.0] *vs*. 75.3 [51.6-109.8] events per 100 person-weeks of observation in PP and control group, respectively), accounting for an HR of 0.84 (95% CI 0.46-1.52; P=0.558). Moreover, no difference in time to hospital discharge was found between the two groups (36.5 [95% CI 23.8-55.9] events per 100 person-weeks of observation among the prone positioning group *vs*. 46.7 [32.0-68.0], among the controls; HR=0.78; 95%CI=0.44-1.38; P=0.397). Adjustment for possible other predictors of clinical progression, such as age and baseline PaO_2_/FiO_2_ ratio, did not change the results to a significant extent.

Using a linear mixed model, change in PaO_2_/FiO_2_ ratio (worst value measured on each day of follow-up) was compared between patients randomized to PP and those in the control group (**Figure 4)**. In both group we observed, on average, an initial decline of the ratio during the first days of observation, followed by an improvement on day 7, which resulted to be slightly higher for the patients randomized to the control group (P=0.042 for the interaction term between time and study arm).

**Figure 4.**
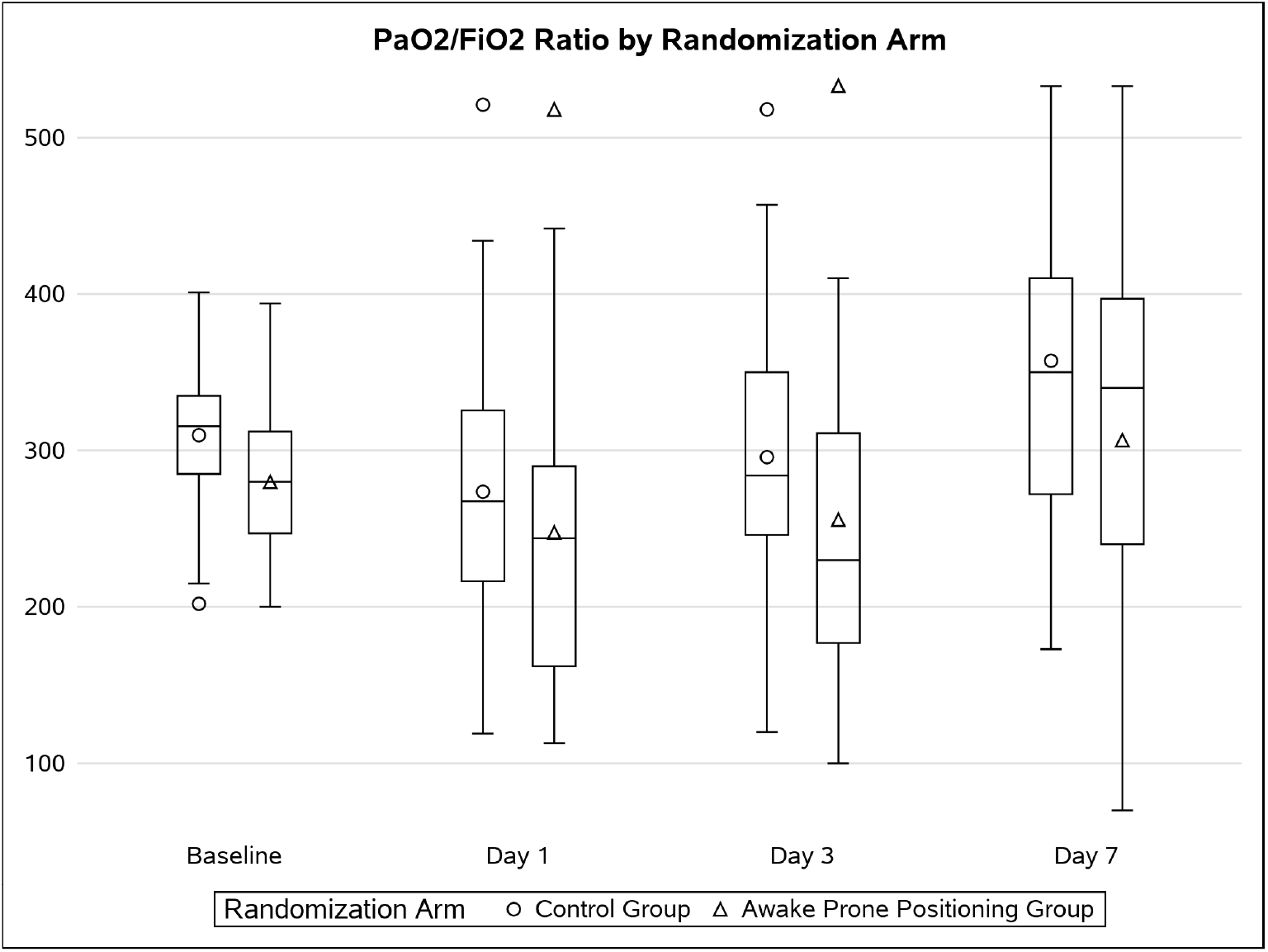
Arterial oxygen partial pressure to fractional inspired oxygen ratio (PaO2/FiO2) over time, by randomization arm. Horizontal lines represent medians, boxes represent interquartile ranges, whiskers represent minimum and maximum values (excluding outliers), the markers represent means and outliers.

### As-treated analysis

Given the suboptimal adherence in a significant subgroup of patients randomized to PP, we conducted an as-treated analysis, in which we included in the intervention group only patients who maintained prone positioning for ≥3 hours/day. In this analysis, which assigned 22 patients to the intervention group and 39 to the control group, the primary outcome was met by 11/22 (50%) patients in the prone positioning group and in 13/39 (33.3%) in the control group (**Figure 3C**). No statistically significant differences were found comparing the incidence of treatment failure in the two groups (23.3 [95%CI 12.9-42.3] *vs*. 13.2 [7.7-22.7] events per 100 person-weeks of observation among PP and controls, respectively, accounting for a HR of 1.8; 95%CI 0.8-3.9; P=0.165). Similarly, using the as-treated approach, no significant differences were found between PP and control group in terms of time to hospital discharge (HR=0.7; 95%CI 0.4-1.3; P=0.300) or oxygen weaning (HR=0.8; 95%CI 0.4-1.6; P=0.582). (**Figure 3D**)

### Safety

No serious adverse events deemed to be caused by the study procedures were reported. Thirteen patients reported at least one adverse event among a pre-defined list of relevant clinical events, including thrombosis or thromboembolism, gastrointestinal bleeding, cardiovascular event, renal failure, diabetes, infections and delirium (**Table 2**). Among them, the most commonly reported were infections (7/61 patients, 11.5%) and venous thromboembolisms (5/61 patients, 8.2%). The rate of these events was comparable between patients randomized to PP and those to standard of care (P=0.544). Ten patients presented an adverse event or a laboratory abnormality (other than those correlated with respiratory failure) graded ≥3 on a 5-grade scale of severity. No statistically significant difference was found according to treatment group (P=0.682)

**Table 2.** Adverse events.

|  |  | Prone positioning<br>group<br>(N=29) | Control group<br>(N=32) |
| --- | --- | --- | --- |
| Clinically relevant<br>adverse events – no. of<br>patients (%) | Thromboembolic event | 2 (6.9%) | 3 (9.4%) |
|  | Gastrointestinal bleeding | 0 | 1 (3.1%) |
|  | Cardiovascular events | 2 (6.9%) | 1 (3.1%) |
|  | Infection | 3 (10.3%) | 4 (12.5%) |
| Grade ≥3 adverse events<sup>a</sup> – no. of patients (%) |  | 4 (13.8%) | 6 (18.7%) |

<sup>a</sup> Severity scale: 1=mild, 2=moderate, 3=serious, 4=life-threatening, 5=fatal.

## DISCUSSION

In this pragmatic, open-label, randomised controlled trial, we found that early PP had no benefit over standard of care in terms of survival, need for respiratory support, oxygen weaning or hospital discharge, among patient hospitalized with mild COVID-19 pneumonia. By contrast, our data suggested a potential harmful effect associated with this procedure.

A previous randomized trial enrolling 400 patients hospitalized with acute respiratory failure due to COVID-19, the majority of whom treated with high-flow oxygen or non-invasive ventilation, failed to demonstrate a statistically significant effect of PP on patient mortality or need for mechanical ventilation, although a numerically lower number of patients treated with PP underwent endotracheal intubation [14]. A subsequent meta-analysis of six randomised controlled open-label trials suggested that awake prone positioning may result in a decreased risk of intubation in patients with COVID-19 who require support with high-flow nasal cannula [18]. Our data did not confirm these findings in a population of patients with less severe respiratory impairment at baseline and even suggested that too early PP could be associated with an increased risk of clinical progression. Consistently with our findings, in a non-randomized controlled trial including mostly patients receiving low-flow oxygen through nasal cannula, those adjudicated to prone positioning had a worse clinical status at 5 days than controls [16]. Moreover, in a randomized trial, conducted in patients using high-flow nasal oxygen or noninvasive ventilation for respiratory support, PP did not reduce the rate of intubation [17]. Eventually, several observational studies showed no beneficial effects on mortality or orotracheal intubation in patients undergoing awake prone positioning [7,19,20].

These apparent contradictions possibly reflect a lack of effectiveness of prone positioning in patients with mild-to-moderate respiratory impairment. Of note, among mechanically ventilated patients, a clear clinical benefit of PP has been demonstrated only for those who require elevated PEEP and have severe ARDS (ie, PaO_2_/ FiO_2_ ratio <150 mmHg) [4,22]. Timing of PP, therefore, may be relevant, because it has been speculated the existence of different phases and phenotypes of ARDS, that may influence its response to pronation [23]. It has been suggested that the early phase of COVID-related ARDS is characterised by high lung compliance [24], a condition that is likely not to benefit from prone positioning because it is characterized by a low amount of non-aerated lung tissue and, consequently, a reduce alveolar recruitability through prone positioning [25]. Taken all together, these findings suggest that awake prone-positioning could be futile in populations like ours, with mild COVID-related pneumonia, and that it should be reserved to patients with more advanced disease, who require a high fraction of inspired oxygen or non-invasive ventilation. On the other hand, however, when PP has been used as a rescue therapy to avoid mechanical ventilation in patient with refractory hypoxemia despite HFNC or NIV, it did not help to reduce the rate of oro-tracheal intubation [17]. Thus, the optimal timing for awake PP is yet to be identified, because an excessive delay may limit its usefulness. It is reasonable to think that a trial of awake prone positioning could be tempted in patients who require HFNC or NIV, but not earlier, and that, in any case, awake PP should not be regarded as a substitute for intubation in patients who meet the indications for mechanical ventilation.

Another possible explanation for our findings could lay in the short time spent by the patients in PP. Although the optimal duration of awake PP has not been established, a recent systematic review suggested that a threshold of 4 hours of pronation per day was adequate to observe a beneficial effect on patient gas exchanges [26]. The scheduled and actual time spent in PP by our patients was close to this threshold and comparable to that of other studies [18,22,24]. Nonetheless, such time was lower than expected, basing on the recommendation given to the patients, and lower than the time generally recommended in mechanically ventilated patients, for whom the time suggested for PP is 12-16 hours per cycle [14]. Although reduced adherence to PP could have hampered its effect, no statistically nor clinically significant benefit was associated with awake pronation even when the analysis was focused only on patients who were actually able to comply with PP reccomendations. Whether maintaining prone position for longer time than that proposed in our study (6 hours/day) may be associated with additional benefit among non-intubated patients merits to be investigated further. Additional efforts to improve patient comfort during pronation and their adherence to the procedure should be pursued.

The reasons why awake PP appeared to be even detrimental, in our study, are difficult to ascertain. Overall, PP was well tolerated while adverse events attributable to PP were rare and not significantly different in the study groups. We cannot exclude, however, that transient improvements in peripheral oxygenation or respiratory rate during pronation could have delayed appropriate therapeutic interventions in patients whose lung function showed signs of deterioration. On the one hand, previous studies showed that improved oxygenation and reduced respiratory rate were observed during prone positioning. However, these changes were transient in most of the cases, as conditions reverted to baseline once the patient returned to the supine position [7,27,28]. On the other hand, delayed access to intubation or non-invasive ventilation could increase the risk of self-induced lung injury due to vigorous respiratory efforts during spontaneous breathing [29,30].

Interestingly enough, we did not observe any beneficial effect of awake PP on gas exchanges, as measured by PaO_2_/ FiO_2_ ratios, in apparent contradiction with previous studies [7]. It should be noted, however, that in our study we reported the lowest PaO_2_/FiO_2_ ratios measured on each day of follow-up, while other studies considered the oxygenation improvement during or at the end of pronation. This suggests that the improvement of gas exchange after prone positioning is likely to be temporary and lost once the patient re-supinates, as other authors have speculated before [27].

Our study has strengths, but also limitations, that merit to be acknowledged. The randomized design allowed to reduce the risk of bias and to obtain comparable groups. Its pragmatic nature reflected the real-world challenges of implementing awake prone positioning. In addition, the selected outcomes (death, use of MV or NIV, oxygen weaning and hospital length of stay) appear as clinically relevant compared to surrogate endpoints such as changes in respiratory parameters. Lastly, the enrolment of patients with mild Covid-19 pneumonia allowed the study of a population less explored in the previous trials on the same subject. However, the study presents also important limitations. First, the unblinded nature of the trial, which was inevitable given the nature of the intervention; second, the small sample size which may reduce the generalizability of our results; third, the suboptimal adherence to awake prone positioning, which calls for the implementation of further studies regarding strategies to increase the time spent in prone positioning.

In conclusion, we observed no evidence of any clinically significant effects of prone positioning in awake patients with mild Covid-19 pneumonia; on the contrary, we can’t rule out that too early or inappropriate PP may cause harm to patients. Thus, awake prone positioning should not be recommended as a routine treatment for COVID-19, at least in patients without severe hypoxemic respiratory failure, out of clinical research aimed at investigating if and in which population it could be more beneficial.

## Data Availability

All data produced in the present study are available upon reasonable request to the authors

## ACKNOWLEDGMENTS

The EPCoT Study Groups includes, besides the authors, also (in alphabetical order) Anna Cappelletti, Ilaria Caramma, Viola Cogliandro, Ilaria De Benedetto, Giulia Gustinetti, Marta Iannace, Valentina Orsini, Alice Ranzani, Anna Spolti.

We kindly thank Anna Coppo and Giacomo Bellani for sharing with us their experience and for their useful advices on the implementation of awake prone positioning.

The authors declare that they have no conflict of interests for the present study.

